# An approximate Bayesian approach for estimation of the reproduction number under misreported epidemic data

**DOI:** 10.1101/2021.05.19.21257438

**Authors:** Oswaldo Gressani, Christel Faes, Niel Hens

## Abstract

In epidemic models, the effective reproduction number is of central importance to assess the transmission dynamics of an infectious disease and to orient health intervention strategies. Publicly shared data during an outbreak often suffers from two sources of misreporting (underreporting and delay in reporting) that should not be overlooked when estimating epidemiological parameters. The main statistical challenge in models that intrinsically account for a misreporting process lies in the joint estimation of the time-varying reproduction number and the delay/underreporting parameters. Existing Bayesian approaches typically rely on Markov chain Monte Carlo (MCMC) algorithms that are extremely costly from a computational perspective. We propose a much faster alternative based on Laplacian-P-splines (LPS) that combines Bayesian penalized B-splines for flexible and smooth estimation of the time-varying reproduction number and Laplace approximations to selected posterior distributions for fast computation. Assuming a known generation interval distribution, the incidence at a given calendar time is governed by the epidemic renewal equation and the delay structure is specified through a composite link framework. Laplace approximations to the conditional posterior of the spline vector are obtained from analytical versions of the gradient and Hessian of the log-likelihood, implying a drastic speed-up in the computation of posterior estimates. Furthermore, the proposed LPS approach can be used to obtain point estimates and approximate credible intervals for the delay and reporting probabilities. Simulation of epidemics with different combinations for the underreporting rate and delay structure (one-day, two-day and weekend delays) show that the proposed LPS methodology delivers fast and accurate estimates outperforming existing methods that do not take into account underreporting and delay patterns. Finally, LPS is illustrated on two real case studies of epidemic outbreaks.

## 1 Introduction

In presence of an epidemic outbreak, it is of vital importance to gain insights into the transmissibility of a disease and have a clear understanding of the mechanisms driving the dynamics of infections over time. Real-time information on epidemiological parameters can have a determinant role in orienting public health policies and initiate proactive interventions for disease control and prevention. The effective reproduction number, *R*_*t*_, defined as the average number of secondary infections generated by a primary infected individual in a susceptible population at a calendar time *t >* 0 (Hethcote, 2000; Bettencourt and Ribeiro, 2008) is probably among the most important parameters that permits to gain knowledge on time-dependent variations in the transmission potential (Nishiura and Chowell, 2009). During the last twenty years or so, serious efforts have been invested in the development of sophisticated inferential methods to estimate the time-varying reproduction number, a challenging task as recently recalled and beautifully summarized by Gostic et al. (2020). Among early contributors, one can cite Wallinga and Teunis (2004) who propose a likelihood-based estimation of the effective reproduction number solely based on information provided by the observed epidemic curve. This work was further extended and generalized by Cauchemez et al. (2006) who assume no prior knowledge on the generation interval, i.e. the time elapsed between when a susceptible person becomes infected (infector) and when that individual infects another person (infectee) (Svensson, 2007). Their model captures the pattern of *R*_*t*_ over time by using partial tracing information and Markov chain Monte Carlo (MCMC) for posterior inference.

Delay in reporting and underreporting of incidence data (Lawless, 1994; Cui and Kaldor, 1998; Fraser et al., 2009) add a further layer of difficulty that cannot be ignored when designing a model to estimate the reproduction number, as misreported data alters the true underlying signal of an epidemic curve and hence introduces bias in estimates of *R*_*t*_. The model of Hens et al. (2011) explicitly accounts for underreporting and provides estimates of *R*_*t*_ based on a frequentist likelihood approach that assumes a fixed serial interval distribution (the time elapsed between symptom onset in an infectee and its infector). Azmon et al. (2014) go one step further and use a Bayesian semiparametric approach with penalized radial splines to model *R*_*t*_ accounting simultaneously for underreporting and delay in reporting. They use the renewal equation (Feller, 1941; Fraser, 2007; Wallinga and Lipsitch, 2007; Nouvellet et al., 2018) to establish a link between *R*_*t*_ and daily incidence counts to describe the evolutionary dynamics of an epidemic. When resorting to Bayesian methods for inference in epidemiological models, MCMC sampling practically imposes itself as a default option as it is a deeply routed and versatile tool that is made accessible and implementable by many computer software packages such as WinBUGS (Lunn et al., 2000) or JAGS (Plummer et al., 2003).

Notwithstanding the capacity of MCMC to explore virtually any posterior target distribution, there is often a large computational price to pay accompanied by eventual convergence problems and the systematic necessity to diagnose MCMC samples. To overcome these limitations and get rid of the computational hurdles imposed by MCMC, we propose a completely sampling-free approximate Bayesian inference approach for fast and flexible estimation of the reproduction number *R*_*t*_ in an epidemic model with misreported data. In particular, we revisit the model of Azmon et al. (2014) by using Bayesian P-splines (Eilers and Marx, 1996; Lang and Brezger, 2004) for flexible estimation of the time-varying reproduction number and Laplace approximations (Rue et al., 2009; Gressani and Lambert, 2018, 2021) to the conditional posterior of the latent spline vector related to *R*_*t*_ for fast computation. Our Laplacian-P-splines (LPS) model is based on the following three assumptions: (1) a closed susceptible population (i.e. no imported cases), (2) the generation interval distribution is assumed to be known and (3) an informative prior on the reporting rate is available. A composite link model (Thompson and Baker, 1981; Eilers, 2007) is used to represent the delay process, of which we investigate three possible structures, one-day, two-day and weekend delays. Moreover, we assume that the mean number of new contaminations is driven by the renewal equation, i.e. the product of the effective reproduction number and a discrete convolution between past cases and generation probabilities. Several simulation scenarios show that the proposed methodology gives accurate estimates of the reporting and delay probabilities and is also able to precisely capture the pattern of *R*_*t*_ over the course of an epidemic. Encouraging results are also observed when comparing LPS with the **EpiEstim** package of Cori et al. (2013) which is known for producing robust estimates of *R*_*t*_. The key advantage of our approach is that even though we work from a completely Bayesian perspective, LPS delivers estimates of key epidemiological model parameters in seconds, while several minutes or hours would be needed with MCMC algorithms. This is partly due to the fact that Laplace approximations are based on analytically derived expressions for the gradient and Hessian of the log-likelihood of the model.

The presentation of the LPS methodology for fast inference of *R*_*t*_ under misreported data is structured as follows. In Section 2, the Laplacian-P-splines model is introduced and priors are imposed on the hyperparameters. After summarizing the Bayesian model, we show how the conditional posterior of the spline vector related to *R*_*t*_ is approached with Laplace approximations. Next, posterior inference on reporting and delay probabilities are presented along with the construction of credible intervals for the latent parameters. Section 3 is devoted to a detailed numerical study that assesses the performance of LPS under various epidemic scenarios. In Section 4, we illustrate our new methodology on real datasets and Section 5 concludes the paper with a discussion.

## 2 The Laplacian Bayesian P-spline model

### 2.1 Misreported epidemic data

Let *T >* 0 denote the total number of days of an epidemic and ℳ = {*M*_1_, …, *M*_*T*_} the latent set of contaminations with *M*_*t*_ ∈ ℕ the (unobserved) number of new contaminations on day *t*. We write *p*_*j*_ for the probability that *j* days have passed until occurrence of infection in an infector-infectee pair and denote by **p** = {*p*_1_, …, *p*_*k*_} the generation interval distribution of maximum length *k*, assumed to be known here. Following Azmon et al. (2014), we assume that *M*_*t*_ is Poisson distributed with mean *µ*_*t*_ and probability mass function:

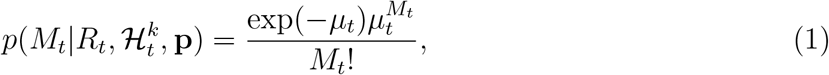

where *R*_*t*_ is the reproduction number at day *t* and 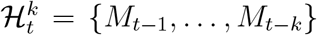 is the set of past values for the number of cases with history of length *k*. The relationship between the mean number of new cases at day *t* and past infections is governed by the epidemic renewal equation:

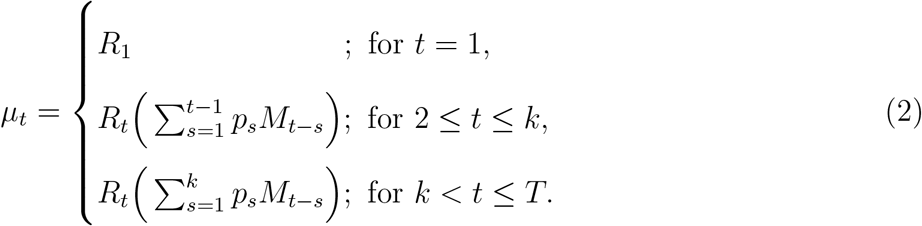

Equation (2) suggests that for *t >* 1, the mean number of new contaminations on day *t* (namely *µ*_*t*_) is a convex combination of the past number(s) in the set 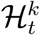 weighted by *R*_*t*_, i.e. the average number of secondary cases generated by a primary case at moment *t*. The observed set of disease counts subject to underreporting and delay in reporting is denoted by 𝒟 = {*O*_1_, …, *O*_*T*_}. The daily reporting probability is given by *ρ* ∈ (0, 1) and is considered to be time-homogeneous over the entire duration of the epidemic. The fraction of cases on day *i* reported on day *t* is written as *δ*_*i→t*_ ∈ [0, 1] and represents a delay probability that can be embedded under various structures in the model via a composite link framework (Eilers, 2007). In this paper, we consider three delay structures proposed in Section 2.2 of Azmon et al. (2014), namely a one-day, two-day and weekend delay pattern. The underreporting-delay process is reflected in the Poisson distributional assumption for *O*_*t*_ with mean 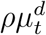:

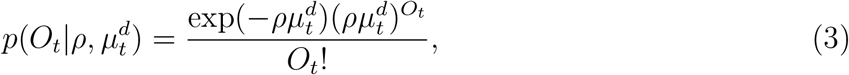

where 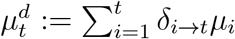is the average number of cases on day *t* subject to delays computed by aggregating the current and past (unobserved) mean number of cases weighted by their associated delay probability. Mathematically, the delay pattern is determined by a square composition matrix 𝒞 of dimension 7 × 7, with rows and columns representing the days of a week. The link between ***µ*** = (*µ*_1_, …, *µ*_*t*_)^⊤^ and 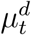 can thus be written compactly as 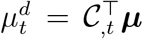, where 𝒞_,*t*_ is column *t* of the composition matrix. Appendix A contains the composition matrices for the three delay patterns considered in this paper.

### 2.2 Bayesian model formulation

#### 2.2.1 Flexible specification of the effective reproduction number

P-splines (Eilers and Marx, 1996) are an interesting candidate to model the time-varying reproduction number dynamics over the considered epidemic period. Two main appealing features of this spline smoother are worth mentioning. First, the penalty matrix can be straightforwardly obtained with minimal numerical effort for any chosen penalty order. Second, P-splines are naturally translated in a Bayesian framework by replacing the deterministic discrete difference penalty by random walks with Gaussian errors (Lang and Brezger, 2004), yielding Gaussian priors for the B-spline coefficients. This motivates our choice for modeling the log of the effective reproduction number as a linear combination of B-splines, i.e. 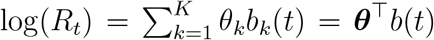, where ***θ*** = (*θ*_1_, …, *θ*_*K*_)^⊤^ is the latent vector of B-spline coefficients and *b*(·) = (*b*_1_(·), …, *b*_*K*_(·))^⊤^ is a basis of cubic B-splines on the domain [0, *T*] ranging from 0 to the last day of the epidemic. A “large” number *K* of B-spline basis functions is specified to ensure that the fitted curve for *R*_*t*_ is flexible enough and a discrete penalty term *λ****θ***^⊤^*P* ***θ*** is introduced as a measure of roughness of the B-spline coefficients with *λ >* 0 as a tuning parameter. The penalty matrix is given by 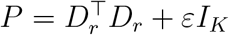 and equals the product of *r*th order difference matrices *D*_*r*_ with a small perturbation on the main diagonal (here *ε* = 10^−5^) to ensure full rankedness. The Gaussian prior for the spline vector is denoted by 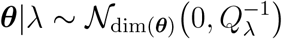, with precision matrix *Q*_*λ*_ = *λP*. The tuning parameter is assigned a Gamma prior *λ* ∼ 𝒢(*a*_*λ*_, *b*_*λ*_) with mean *a*_*λ*_*/b*_*λ*_ and variance 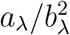. Choosing *a*_*λ*_ = *b*_*λ*_ = 10^−5^ yields a dispersed (yet proper) prior for *λ* with a large variance (see e.g. Lang and Brezger, 2004; Lambert and Eilers, 2005).

#### 2.2.2 Prior assumptions on reporting and delay probabilities

The tuning parameter and the reporting and delay probabilities are gathered in the hyperparameter vector ***η*** = (*λ, ρ, δ*_*i→j*_; *i, j* = 1, …, 7)^⊤^. A noninformative uniform prior is imposed on the delay probabilities, i.e. *δ*_*i→j*_ ∼ 𝒰(0, 1) for *i, j* = 1, …, 7. Typically, the reporting rate *ρ* cannot be extrapolated from real-time data (Heesterbeek et al., 2015) and thus needs to be estimated, adding an extra layer of difficulty in the inference process. As noted by Thompson et al. (2019), underreporting has already proved to be a burden for inference and forecasting in various infectious disease models. Without minimal prior knowledge on *ρ*, posterior estimates of key epidemiological quantities such as the effective reproduction number will likely be biased and accompanied by high uncertainty with wide credible intervals. We therefore assume that minimal prior information is available for the reporting probability translated by a uniform prior *ρ* ∼ 𝒰(*a*_*ρ*_, *b*_*ρ*_) with bounds 0 *< a*_*ρ*_ *< b*_*ρ*_ *<* 1 that encompass the true underlying *ρ*. Such informative priors can for instance be constructed using hierarchical models based on available historical data (Riou et al., 2018) or by using posterior distributions of *ρ* inferred from previous studies (Stocks et al., 2020).

To approximate the (latent) number of cases on a given day *M*_*t*_, we use a simple inflation factor approach as in Jandarov et al. (2014) and Stocks et al. (2020) based on our prior assumption for *ρ*. More specifically, we use the following approximation 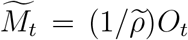, where 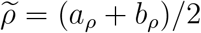 is the midpoint in the prior domain [*a*_*ρ*_, *b*_*ρ*_]. Although more sophisticated methods exist to account for underreporting (see e.g. Bracher and Held, 2020), the main rationale for using a simple multiplication rule is that the focus of this paper is on the methodological approach for fast approximate Bayesian inference of *R*_*t*_ taking reporting/delay into account, rather than on an explicit modeling of the (under)reporting process in itself. To summarize, the full Bayesian model is given by:

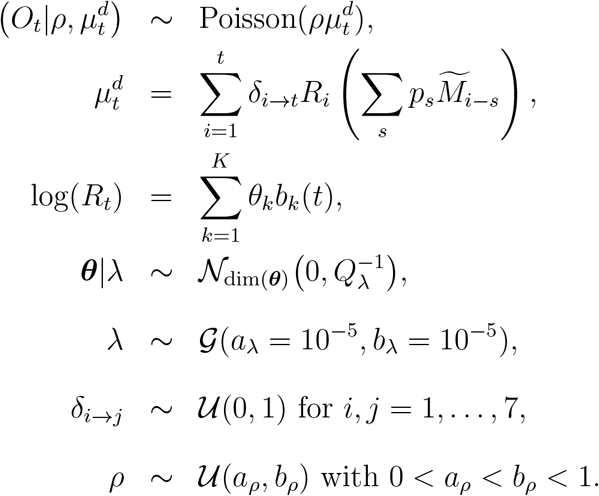

### 2.3 Approximation of the conditional posterior spline vector

The log-likelihood function of the Poisson model for the observed number of cases is:

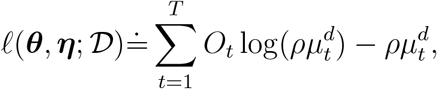

where ≐ denotes equality up to an additive constant. Replacing 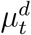 by its extensive form in terms of the epidemic renewal equation and the spline specification of the effective reproduction number, the mean of *O*_*t*_, namely 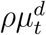, is written as the following function:

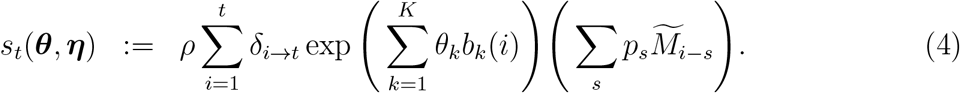

The above equation is used to write the log-likelihood as follows:

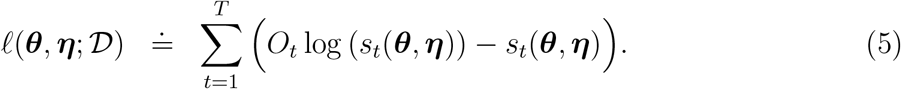

Using (5) and Bayes’ rule, the conditional posterior of the spline vector is:

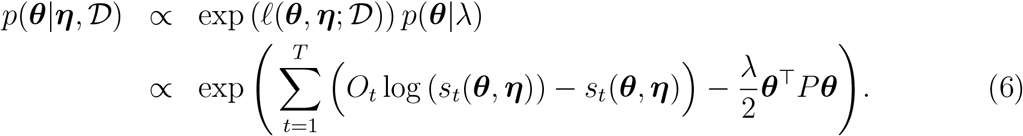

Let *g*_*t*_(***θ, η***) := *O*_*t*_ log(*s*_*t*_(***θ, η***)) − *s*_*t*_(***θ, η***) denote the contribution of observables at day *t* to the log-likelihood. A Laplace approximation to *p*(***θ***|***η***, 𝒟) is obtained by iteratively computing a second-order Taylor expansion of *g*_*t*_(***θ, η***) in terms of ***θ*** by starting from an initial guess ***θ***^(0)^. The Taylor expansion to *g*_*t*_(***θ, η***) yields a quadratic form in ***θ*** and plugging the latter into (6), one recovers (up to a multiplicative constant) a multivariate Gaussian density. The iterative Laplace approximation scheme is implemented in a Newton-Raphson type algorithm for which the gradient and Hessian of *g*_*t*_(***θ, η***) and hence of the log-likelihood are analytically derived in Appendix B for maximum numerical efficiency. The Laplace approximated conditional posterior of the B-spline vector after convergence of the Newton-Raphson algorithm is denoted by 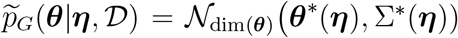, where ***θ***^*^(***η***) is the mean (mode) and Σ^*^(***η***) the variance-covariance matrix, for a given value of the hyperparameter vector ***η***.

### 2.4 Posterior inference on hyperparameters

The posterior of the hyperparameter vector ***η*** can be written in terms of the conditional posterior of the B-spline vector derived in Section 2.3, namely:

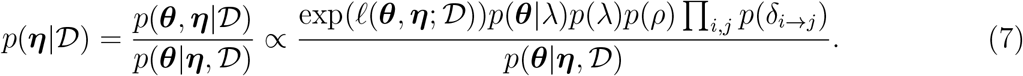

Following Tierney and Kadane (1986) and Rue et al. (2009), the above hyperparameter posterior can be approximated by replacing the denominator in (7) with its Laplace approximation and by substituting ***θ*** by the modal value ***θ***^*^(***η***) of the latter Laplace approximation. The resulting approximated posterior is then solely a function of ***η***:

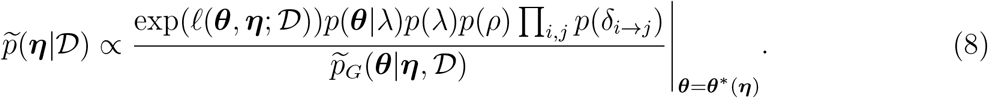

The uniform priors on the reporting and delay probabilities vanish into the proportionality constant and so the approximated hyperparameter posterior (8) is written extensively as:

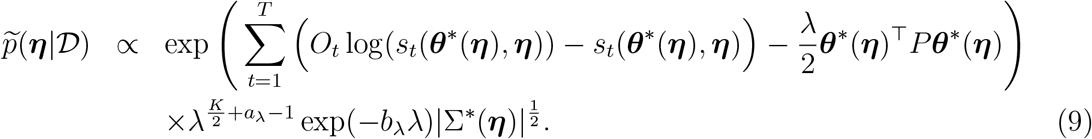

As the hyperparameters live in different domains, e.g. *λ >* 0 and *ρ* ∈ (0, 1), we propose a transformation to ensure that all variables are unbounded with values in ℝ. This transformation is crucial to ensure numerical stability when using algorithms to explore 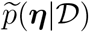. Let us define *v* = log(*λ*), 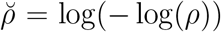and 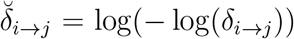 for *i, j* = 1, …, 7 and denote our transformed hyperparameter vector as 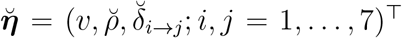. Using the multivariate transformation method, the hyperparameter posterior becomes:

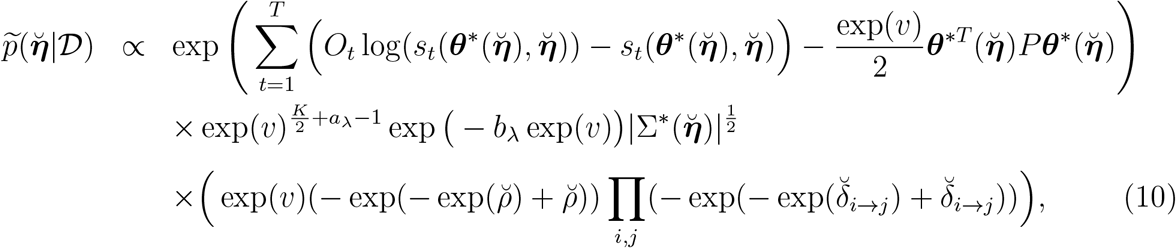

where the last line equals the absolute value of the Jacobian from the multivariate transformation. The approximate posterior of the transformed hyperparameter vector in (10) is the main ingredient for posterior inference on ***η***. At this stage, MCMC methods could be used to explore the above (approximate) target density (see e.g. Vanhatalo et al., 2013; Gómez-Rubio and Rue, 2018). As the philosophy of our approach is to rely on a completely sampling free methodology, we decide to compute the maximum a posteriori (MAP) estimate of 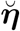 via a Newton-Raphson algorithm. Even though MAP approaches ignore the uncertainty surrounding the estimate contrary to grid-based strategies or MCMC samplers, they have the advantage of being less costly to implement from a computational perspective and still have good statistical properties as will be shown later in the simulation study.

When designing a Newton-Raphson algorithm to explore a complex posterior as in (10), great care needs to be taken to avoid convergence problems. For maximization, it is important to ensure that an ascent direction is taken at every iteration. This can be achieved by proposing a modified positive definite version of the negative Hessian whenever the latter fails to be positive definite (Levenberg, 1944; Marquardt, 1963; Goldfeld et al., 1966) combined with a backtracking strategy (e.g. step-halving) to ensure heading uphill.

### 2.5 Approximate posterior estimates and credible intervals

Let us denote by 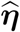 the MAP estimate of ***η*** obtained from the Newton-Raphson algorithm. Plugging the latter into the Laplace approximation scheme, one recovers 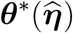 and 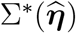, i.e. the point estimate of the B-spline vector and its associated estimated variance-covariance matrix. Using the latter quantities, a point estimate of the effective reproduction number is taken to be 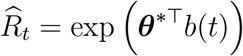. Let *h*(***θ***|*t*) = log(*R*_*t*_) = ***θ***^⊤^*b*(*t*) be the log of the reproduction number at a given day *t* seen as a function of the spline vector ***θ*** and consider the first-order Taylor expansion of *h*(***θ***|*t*) around 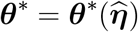:

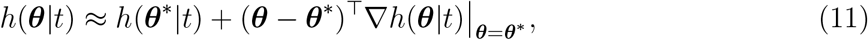

with gradient ∇*h*(***θ***|*t*)|_θ=θ*_ = *b*(*t*). Note that (11) is a linear combination of the random vector ***θ*** and that the latter has a Gaussian (conditional) posterior due to the Laplace approximation scheme. It follows that a posteriori *h*(***θ***|*t*) is also approximately Gaussian with mean 𝔼(*h*(***θ***|*t*)) ≈ *h*(***θ***^*^|*t*) and covariance matrix 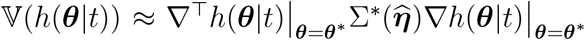. Accordingly, a (1 − *α*) × 100% (approximate) quantile-based credible interval for log(*R*_*t*_) on day *t* is:

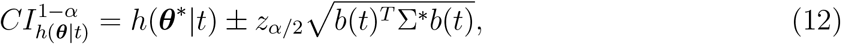

where *z*_*α/*2_ is the *α/*2-upper quantile of a standard normal distribution. Applying the exp(·) transform on (12) yields the desired credible interval for *R*_*t*_.

For the hyperparameter vector, we advise the use of a heavier tailed distribution and assume that the marginal posterior of a hyperparameter variable 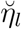 has a Student-t distribution (see e.g. Martins and Rue, 2014) with *ν* = dim(***η***) − 5 degrees of freedom, namely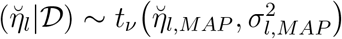, where the mean 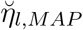 is the MAP estimate from the Newton-Raphson algorithm and 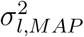 is the appropriate diagonal entry of the inverse of the negative Hessian of the log of (10) evaluated at the MAP estimate. The resulting (1 − *α*) × 100% credible interval for 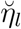 is then 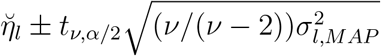.

## 3 Numerical study

The performance of our approach (with cubic B-splines and a penalty of order 3) is assessed in five different epidemic scenarios with a duration of *T* = 30 days. In Scenarios 1-3, the incidence data is governed by a decaying effective reproduction number *R*_*t*_ = exp(cos(*t/*13)) as in Azmon et al. (2014) with a reporting rate fixed at *ρ* = 0.2. Scenarios 4 and 5 assume more complex structures for *R*_*t*_ to see whether the Laplacian-P-splines model is able to capture the true dynamics of the reproduction number. Table 1 summarizes the functional form of the reproduction number, the delay structure, the reporting rate and the assumed prior information on *ρ* used in each scenario. We simulate *S* = 500 replications in each scenario assuming an epidemic starting with *M*_1_ ∼ Poisson(*R*_1_) cases on day *t* = 1 and a generation interval distribution of length *k* = 3 with **p** = {0.2, 0.5, 0.3}. Figure 1 shows the latent (*M*_*t*_ in red) and observed (*O*_*t*_ in orange) incidence data for Scenarios 1-3.

**Table 1:** Functional form of the reproduction number, delay structure, reporting rate and informative prior on *ρ* for the five different simulation scenarios.

| Scenario | Reproduction number | Delay | $\rho$ | Prior for $\rho$ |
| --- | --- | --- | --- | --- |
| 1 | $R_t = \exp(\cos(t/13))$ | One-day | $\rho = 0.20$ | $\rho \sim \mathcal{U}(0, 0.40)$ |
| 2 | $R_t = \exp(\cos(t/13))$ | Two-day | $\rho = 0.20$ | $\rho \sim \mathcal{U}(0, 0.40)$ |
| 3 | $R_t = \exp(\cos(t/13))$ | Weekend | $\rho = 0.20$ | $\rho \sim \mathcal{U}(0, 0.40)$ |
| 4 | $R_t = \exp(0.40 + 0.30 \cos(t/10) + 0.80 \sin((t\pi)/6))$ | One-day | $\rho = 0.60$ | $\rho \sim \mathcal{U}(0.25, 0.85)$ |
| 5 | $R_t = \exp(0.98 + \sin((1.75 + 0.022t)^2)) - 0.10$ | Two-day | $\rho = 0.70$ | $\rho \sim \mathcal{U}(0.34, 0.94)$ |

**Figure 1:**
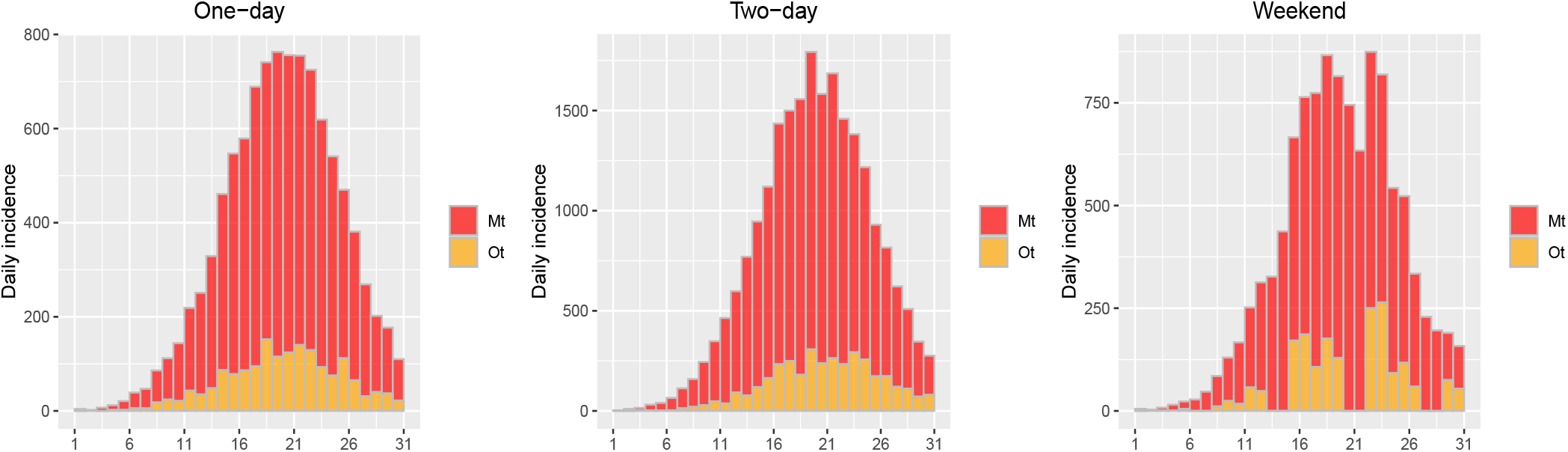
Stacked histograms of daily incidence for different delay patterns (Scenarios 1-3).

For each scenario, we report the mean estimate, empirical standard error (ESE), root mean square error (RMSE) and coverage probability for a 95% credible interval on the hyperparameters. The performance metric for *R*_*t*_ is taken to be the Mean Absolute Error (MAE) defined as 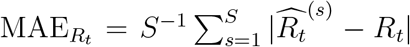. We also compare how LPS performs against the `estimate_R()` function of the **EpiEstim** package (Cori et al., 2013). In particular, we use the syntax `estimate_R(incid=Mtestim, method=“non_parametric_si”, config = make_config(list(si_distr=c(0,p))))`, where `Mtestim` corresponds to the (inflated) number of contaminations. Similarly, we use `incid=Observed` to obtain estimates based on the observed number of cases without an inflation factor, which is most commonly used in the literature. We choose the method `“non_parametric_si”` to specify the distribution of the serial interval, or as is the case here, the generation interval distribution denoted by `p` and inject the latter in the `make_config` option. Moreover, we keep the default option that estimates *R*_*t*_ on weekly sliding windows and use the posterior mean as a point estimate.

Table 2 summarizes the results related to the hyperparameter estimates for Scenarios 1-3 and the left column of Figure 2 shows the estimated curves for *R*_*t*_ (gray) and the pointwise median (in red) of the *S* = 500 estimated curves obtained with LPS. Figure 2 right column shows the MAE of *R*_*t*_ for days *t* = 8, …, 30 obtained with LPS (green) and the **EpiEstim** package with inflation factor on contaminations (light blue) and without inflation factor (dark blue). Estimates of the delay probabilities are relatively close to their true value with a coverage probability slightly above the 95% nominal value. Mean estimates of the reporting probability are close to the true value (*ρ* = 0.2) in all scenarios with a more pronounced undercoverage under a two-day delay pattern (as in Azmon et al., 2014). It is also worth mentioning that the downward trend of *R*_*t*_ is well captured under the three considered delay patterns. Furthermore, Figure 2 (right column), shows that LPS is competitive against **EpiEstim** regarding the estimation of *R*_*t*_.

**Table 2:** Simulation results for the reporting and delay probabilities under Scenarios 1-3 with *R*_*t*_ = exp(cos(*t/*13)), *ρ* = 0.2 and *S* = 500.

| Delay pattern | Parameter | True value | Mean | ESE | RMSE | CI.low | CI.up | CP95% |
| --- | --- | --- | --- | --- | --- | --- | --- | --- |
| One-day<br>(Scenario 1) | $\delta_{Mo \rightarrow Tu}$ | 0.4 | 0.294 | 0.103 | 0.148 | 0.025 | 0.694 | 100.0 |
| | $\delta_{Tu \rightarrow We}$ | 0.5 | 0.360 | 0.088 | 0.165 | 0.060 | 0.704 | 99.6 |
| | $\delta_{We \rightarrow Th}$ | 0.7 | 0.595 | 0.087 | 0.136 | 0.175 | 0.851 | 99.0 |
| | $\delta_{Th \rightarrow Fr}$ | 0.3 | 0.196 | 0.088 | 0.136 | 0.008 | 0.625 | 100.0 |
| | $\delta_{Fr \rightarrow Sa}$ | 0.4 | 0.253 | 0.093 | 0.174 | 0.020 | 0.643 | 100.0 |
| | $\delta_{Sa \rightarrow Su}$ | 0.6 | 0.497 | 0.085 | 0.134 | 0.146 | 0.771 | 99.8 |
| | $\delta_{Su \rightarrow Mo}$ | 0.5 | 0.456 | 0.094 | 0.103 | 0.102 | 0.766 | 100.0 |
| | $\rho$ | 0.2 | 0.203 | 0.013 | 0.013 | 0.179 | 0.227 | 90.6 |
| Two-day<br>(Scenario 2) | $\delta_{Mo \rightarrow Tu}$ | 0.3 | 0.329 | 0.118 | 0.121 | 0.019 | 0.741 | 100.0 |
| | $\delta_{Mo \rightarrow We}$ | 0.4 | 0.332 | 0.134 | 0.150 | 0.049 | 0.686 | 99.8 |
| | $\delta_{Tu \rightarrow We}$ | 0.4 | 0.282 | 0.103 | 0.157 | 0.014 | 0.704 | 99.8 |
| | $\delta_{Tu \rightarrow Th}$ | 0.3 | 0.229 | 0.096 | 0.119 | 0.021 | 0.600 | 99.8 |
| | $\delta_{We \rightarrow Th}$ | 0.3 | 0.212 | 0.079 | 0.118 | 0.006 | 0.635 | 100.0 |
| | $\delta_{We \rightarrow Fr}$ | 0.4 | 0.260 | 0.106 | 0.175 | 0.028 | 0.632 | 99.6 |
| | $\delta_{Th \rightarrow Fr}$ | 0.5 | 0.385 | 0.107 | 0.157 | 0.050 | 0.745 | 100.0 |
| | $\delta_{Th \rightarrow Sa}$ | 0.3 | 0.264 | 0.097 | 0.103 | 0.037 | 0.605 | 100.0 |
| | $\delta_{Fr \rightarrow Sa}$ | 0.3 | 0.203 | 0.085 | 0.129 | 0.005 | 0.626 | 100.0 |
| | $\delta_{Fr \rightarrow Su}$ | 0.4 | 0.307 | 0.103 | 0.139 | 0.054 | 0.639 | 99.6 |
| | $\delta_{Sa \rightarrow Su}$ | 0.5 | 0.404 | 0.124 | 0.156 | 0.038 | 0.782 | 100.0 |
| | $\delta_{Sa \rightarrow Mo}$ | 0.3 | 0.286 | 0.111 | 0.112 | 0.032 | 0.650 | 100.0 |
| | $\delta_{Su \rightarrow Mo}$ | 0.3 | 0.294 | 0.115 | 0.115 | 0.012 | 0.718 | 100.0 |
| | $\delta_{Su \rightarrow Tu}$ | 0.6 | 0.434 | 0.115 | 0.202 | 0.088 | 0.762 | 96.0 |
| | $\rho$ | 0.2 | 0.218 | 0.017 | 0.024 | 0.201 | 0.235 | 45.0 |
| Weekend<br>(Scenario 3) | $\delta_{Mo \rightarrow Tu}$ | 0.4 | 0.362 | 0.152 | 0.156 | 0.000 | 0.904 | 100.0 |
| | $\delta_{Tu \rightarrow We}$ | 0.5 | 0.189 | 0.088 | 0.323 | 0.003 | 0.672 | 98.4 |
| | $\delta_{We \rightarrow Th}$ | 0.7 | 0.494 | 0.074 | 0.218 | 0.163 | 0.759 | 76.0 |
| | $\delta_{Th \rightarrow F}$ | 0.3 | 0.369 | 0.096 | 0.118 | 0.091 | 0.679 | 99.8 |
| | $\delta_{Fr \rightarrow Mo}$ | 0.4 | 0.546 | 0.144 | 0.204 | 0.103 | 0.850 | 100.0 |
| | $\delta_{Sa \rightarrow Mo}$ | 0.6 | 0.576 | 0.136 | 0.138 | 0.000 | 0.968 | 100.0 |
| | $\delta_{Su \rightarrow Mo}$ | 0.5 | 0.439 | 0.115 | 0.130 | 0.000 | 0.954 | 100.0 |
| | $\rho$ | 0.2 | 0.214 | 0.016 | 0.021 | 0.190 | 0.238 | 69.0 |

**Figure 2:**
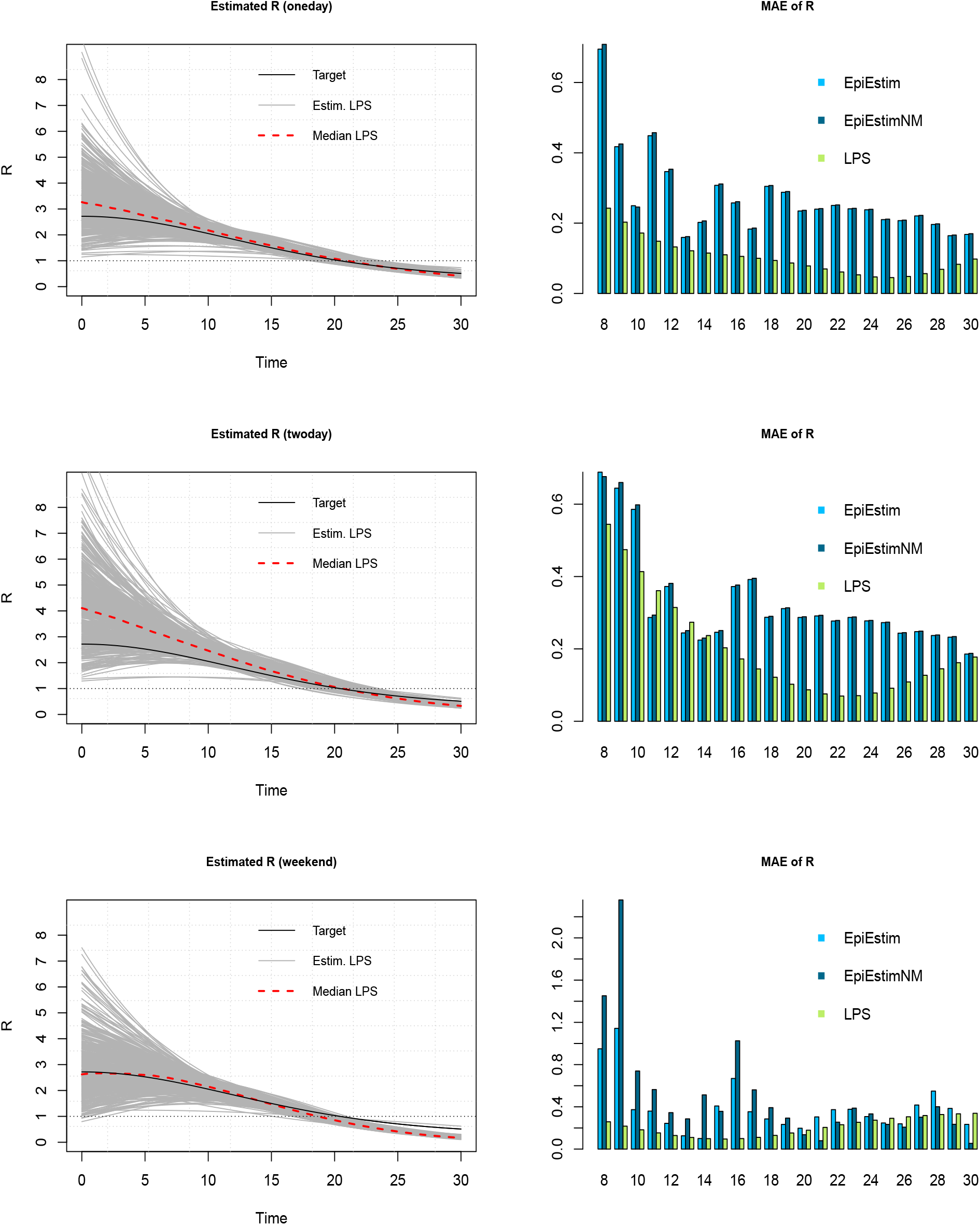
(Left column) Estimation of the time-varying reproduction number (gray curves) and pointwise median (dashed) with LPS for Scenarios 1-3 (rows). (Right column) MAE of *R*_*t*_ for days *t* = 8, …, 30 with LPS (green), EpiEstim (light blue) with multiplication factor and EpiEstimNM (dark blue) ignoring the multiplication factor on contaminations.

Simulation results for the hyperparameters in Scenarios 4 and 5 are given in Table 3. Again, the estimates are relatively close to their true value and the coverages are all reasonable. The undercoverage of certain delay probabilities in Scenario 4 can be explained by the complex shape of *R*_*t*_ and the fact that using a simple inflation factor to recover the latent number of contaminations may be too simplistic here. Figure 3 shows that the estimated *R*_*t*_ obtained with LPS captures the real underlying trend even with more complex structures such as in Scenario 4. In the latter scenario, the MAE of *R*_*t*_ is quite high with **EpiEstim**, while it remains reasonably low with LPS.

**Table 3:**
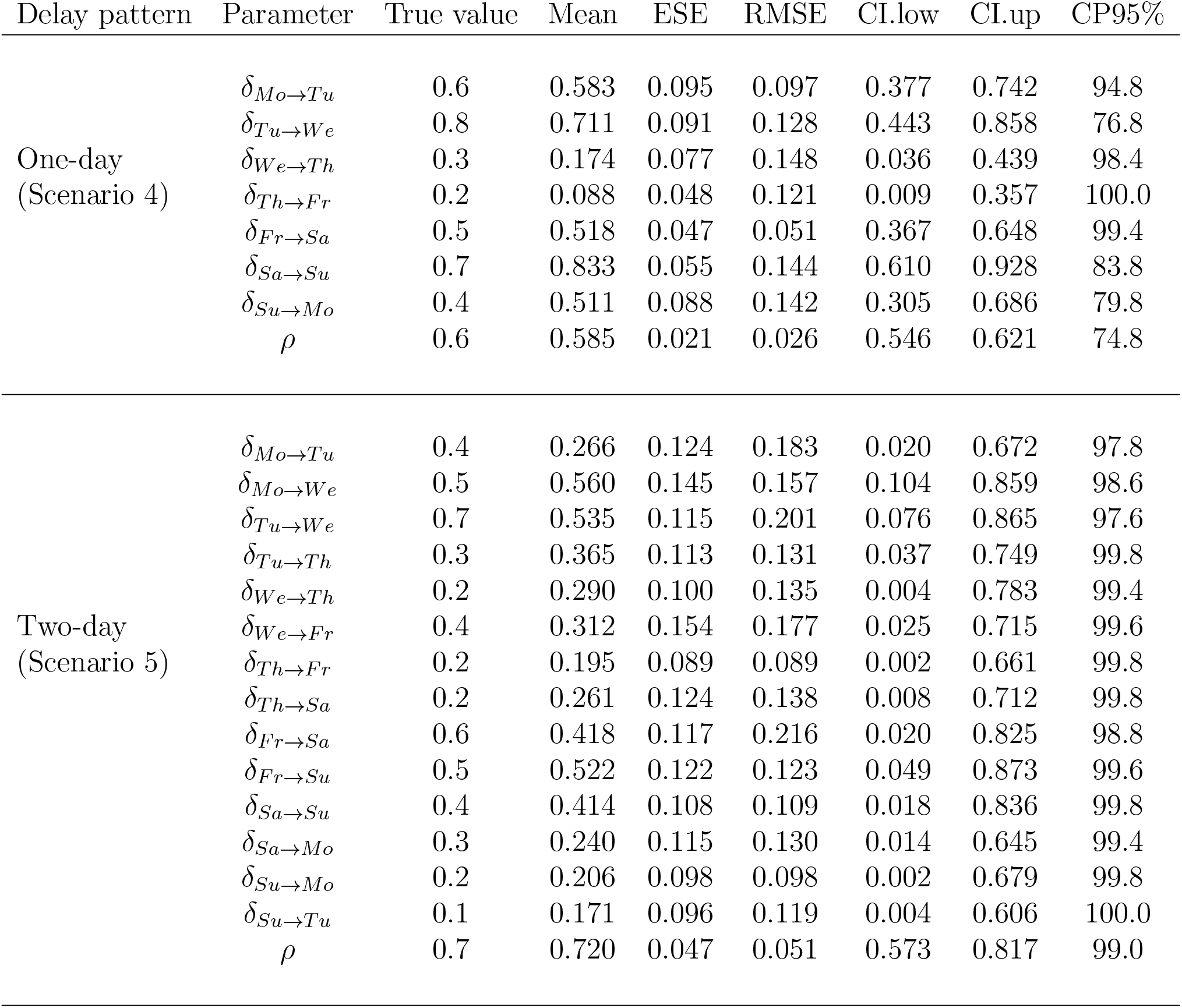
Simulation results for the reporting and delay probabilities under Scenario 4 and 5 with *S* = 500.

| Delay pattern | Parameter | True value | Mean | ESE | RMSE | CI.low | CI.up | CP95% |
| --- | --- | --- | --- | --- | --- | --- | --- | --- |
| One-day<br>(Scenario 4) | $\delta_{Mo \rightarrow Tu}$ | 0.6 | 0.583 | 0.095 | 0.097 | 0.377 | 0.742 | 94.8 |
| | $\delta_{Tu \rightarrow We}$ | 0.8 | 0.711 | 0.091 | 0.128 | 0.443 | 0.858 | 76.8 |
| | $\delta_{We \rightarrow Th}$ | 0.3 | 0.174 | 0.077 | 0.148 | 0.036 | 0.439 | 98.4 |
| | $\delta_{Th \rightarrow Fr}$ | 0.2 | 0.088 | 0.048 | 0.121 | 0.009 | 0.357 | 100.0 |
| | $\delta_{Fr \rightarrow Sa}$ | 0.5 | 0.518 | 0.047 | 0.051 | 0.367 | 0.648 | 99.4 |
| | $\delta_{Sa \rightarrow Su}$ | 0.7 | 0.833 | 0.055 | 0.144 | 0.610 | 0.928 | 83.8 |
| | $\delta_{Su \rightarrow Mo}$ | 0.4 | 0.511 | 0.088 | 0.142 | 0.305 | 0.686 | 79.8 |
| | $\rho$ | 0.6 | 0.585 | 0.021 | 0.026 | 0.546 | 0.621 | 74.8 |
| Two-day<br>(Scenario 5) | $\delta_{Mo \rightarrow Tu}$ | 0.4 | 0.266 | 0.124 | 0.183 | 0.020 | 0.672 | 97.8 |
| | $\delta_{Mo \rightarrow We}$ | 0.5 | 0.560 | 0.145 | 0.157 | 0.104 | 0.859 | 98.6 |
| | $\delta_{Tu \rightarrow We}$ | 0.7 | 0.535 | 0.115 | 0.201 | 0.076 | 0.865 | 97.6 |
| | $\delta_{Tu \rightarrow Th}$ | 0.3 | 0.365 | 0.113 | 0.131 | 0.037 | 0.749 | 99.8 |
| | $\delta_{We \rightarrow Th}$ | 0.2 | 0.290 | 0.100 | 0.135 | 0.004 | 0.783 | 99.4 |
| | $\delta_{We \rightarrow Fr}$ | 0.4 | 0.312 | 0.154 | 0.177 | 0.025 | 0.715 | 99.6 |
| | $\delta_{Th \rightarrow Fr}$ | 0.2 | 0.195 | 0.089 | 0.089 | 0.002 | 0.661 | 99.8 |
| | $\delta_{Th \rightarrow Sa}$ | 0.2 | 0.261 | 0.124 | 0.138 | 0.008 | 0.712 | 99.8 |
| | $\delta_{Fr \rightarrow Sa}$ | 0.6 | 0.418 | 0.117 | 0.216 | 0.020 | 0.825 | 98.8 |
| | $\delta_{Fr \rightarrow Su}$ | 0.5 | 0.522 | 0.122 | 0.123 | 0.049 | 0.873 | 99.6 |
| | $\delta_{Sa \rightarrow Su}$ | 0.4 | 0.414 | 0.108 | 0.109 | 0.018 | 0.836 | 99.8 |
| | $\delta_{Sa \rightarrow Mo}$ | 0.3 | 0.240 | 0.115 | 0.130 | 0.014 | 0.645 | 99.4 |
| | $\delta_{Su \rightarrow Mo}$ | 0.2 | 0.206 | 0.098 | 0.098 | 0.002 | 0.679 | 99.8 |
| | $\delta_{Su \rightarrow Tu}$ | 0.1 | 0.171 | 0.096 | 0.119 | 0.004 | 0.606 | 100.0 |
| | $\rho$ | 0.7 | 0.720 | 0.047 | 0.051 | 0.573 | 0.817 | 99.0 |

**Figure 3:**
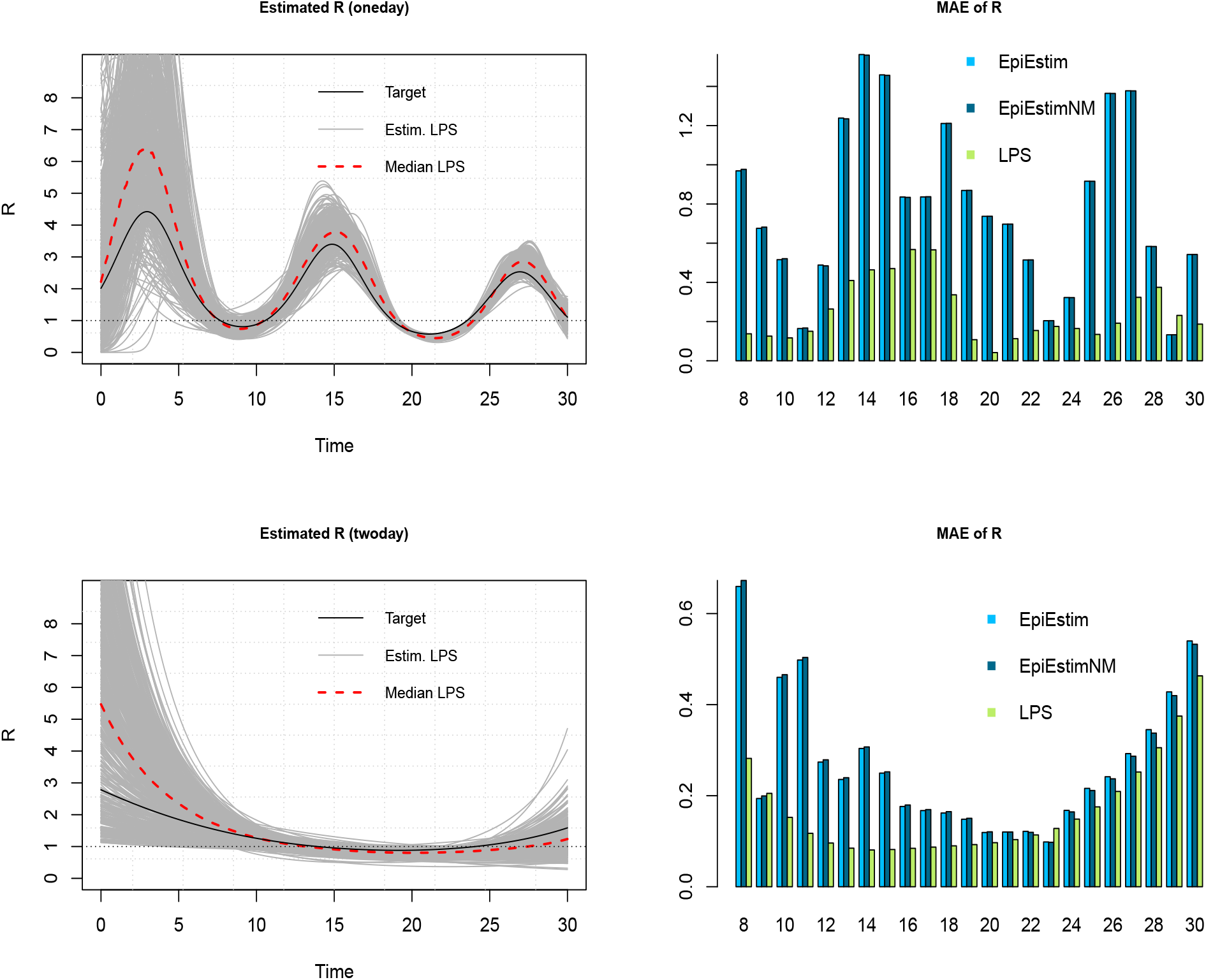
(Left column) Estimation of the time-varying reproduction number (gray curves) and pointwise median (dashed) with LPS for Scenario 4 (top) and 5 (bottom). (Right column) MAE of *R*_*t*_ for days *t* = 8, …, 30 with LPS (green), EpiEstim (light blue) with multiplication factor and EpiEstimNM (dark blue) ignoring the multiplication factor on contaminations.

In terms of computational speed, the LPS approach is extremely fast. With an Intel Xeon E-2186M CPU running at 2.90 GHz, the elapsed real time for fitting the model with misreported data was recorded to be on average 6 seconds for a one-day or weekend delay and a little bit more (around 8 seconds) for a two-day delay as the hyperparameter dimension is larger in the latter case. This is substantially less than any existing MCMC algorithm that typically take minutes (or hours) to fit such complex epidemic models. Furthermore, as LPS does not rely on any sampling scheme, the additional burden of diagnostic checks (e.g. trace plots, Geweke statistics) is also avoided.

## 4 Real data applications

### 4.1 The 1918 influenza pandemic in Baltimore

The LPS methodology is first illustrated in the context of the 1918 H1N1 influenza pandemic in Baltimore, USA with data obtained from the **EpiEstim** package (Cori et al., 2013). The dataset contains daily incidence of onset of disease for a period of 92 days and a discrete daily distribution of the serial interval for influenza. We assume a serial interval of three days and use the latter as a proxy for the generation interval. Based on the serial interval data of the **EpiEstim** package we thus fix **p** = {0.233, 0.359, 0.408} for the generation interval.

As no information is available on the underreporting rate, we use the prior *ρ* ∼ 𝒰(0, 1) without inflating the observed incidence data and assume a one-day delay pattern. For a smooth estimation of the reproduction number, we use *K* = 20 (cubic) B-splines in [0, 92] and a third-order penalty to counterbalance the flexibility of the fitted curve. Figure 4 shows the daily incidence of the 1918 H1N1 data (top) and the estimated time-varying reproduction number (bottom) with LPS (solid), the `estimate_R()` routine of the **EpiEstim** package (dashed) and the Wallinga and Teunis (2004) method (dotted). The gray surface is the (approximate) 95% pointwise credible interval for *R*_*t*_ obtained with LPS. Around day *t* = 30, the estimated *R*_*t*_ reaches a peak before gradually decaying towards one, a pattern also observed in White and Pagano (2008). The reporting rate is estimated to be 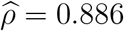 with a 95% credible interval [0.832; 0.923].

**Figure 4:**
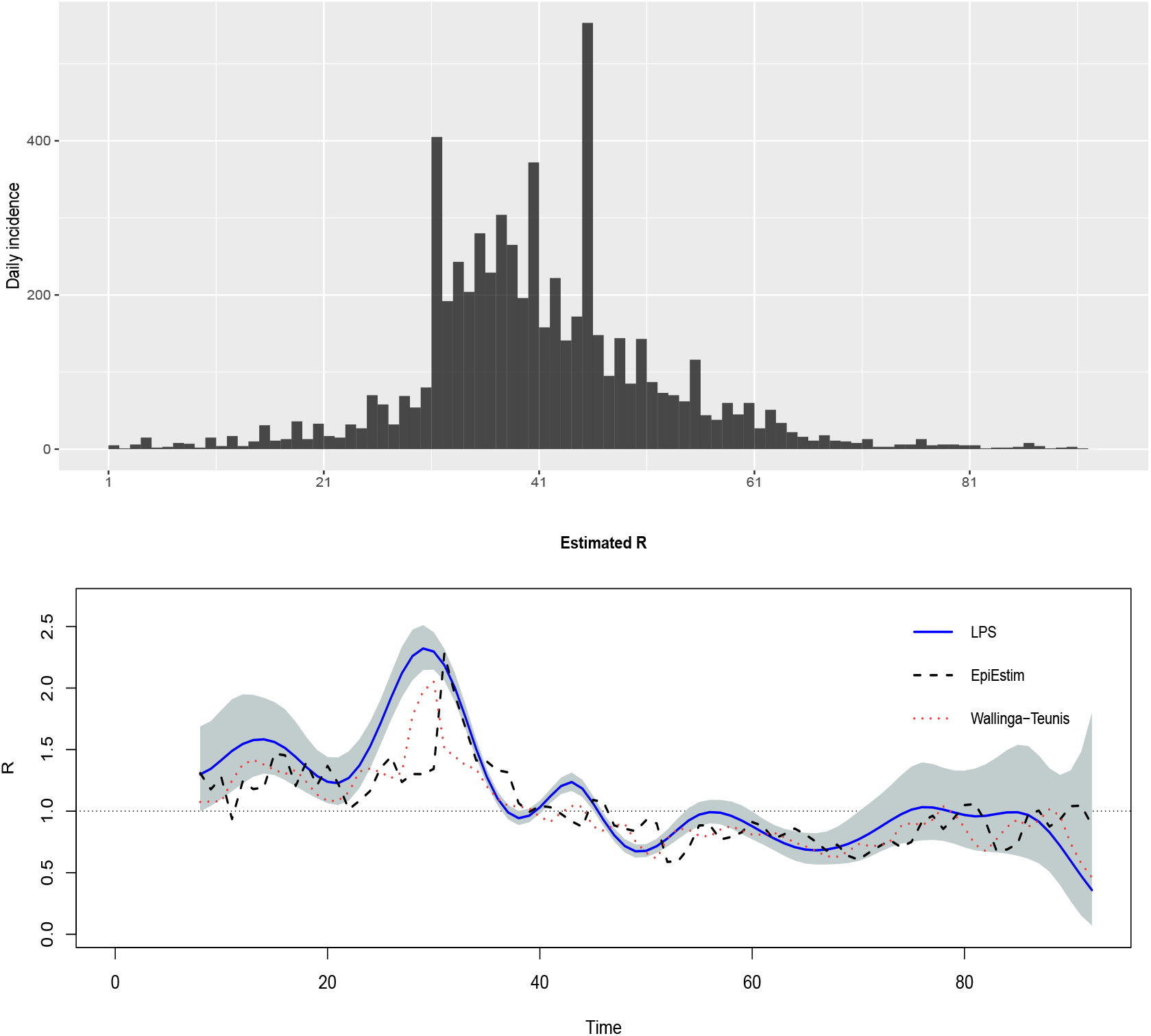
Daily incidence (top) and estimated *R*_*t*_ for the 1918 Influenza data in Baltimore with the **EpiEstim** (dashed), LPS (solid) and Wallinga-Teunis (dotted) method.

### 4.2 Covid-19 data for Australia

In a second application, we use LPS to estimate the time-varying reproduction number of Covid-19 hospitalizations for Australia between May and September 2020. The data is obtained from the **COVID19** package (Guidotti and Ardia, 2020). We use a generation interval of length five, namely **p** = {0.1, 0.2, 0.4, 0.2, 0.1} and estimate the model under a one-day and two-day delay structure with prior *ρ* ∼ 𝒰(0.7, 0.8) and hence an inflation factor of 1*/*0.75 on the observed number of cases.

Figure 5 shows the daily incidence (top) and the estimated *R*_*t*_ under the two considered delay structures. There is a strong similarity between the estimated pattern of *R*_*t*_ for the three considered methods (LPS, EpiEstim and Wallinga-Teunis). Also, comparing the LPS estimate of the reproduction number with the real-time estimates of Arroyo-Marioli et al. (2021) in their online dashboard that uses a Kalman filter as a tool for inference, we also see a similar pattern. In end May, *R*_*t*_ is below one and increases during the next two months to reach a peak in July 2020. Then it slowly decreases to reach a value below one around end August.

**Figure 5:**
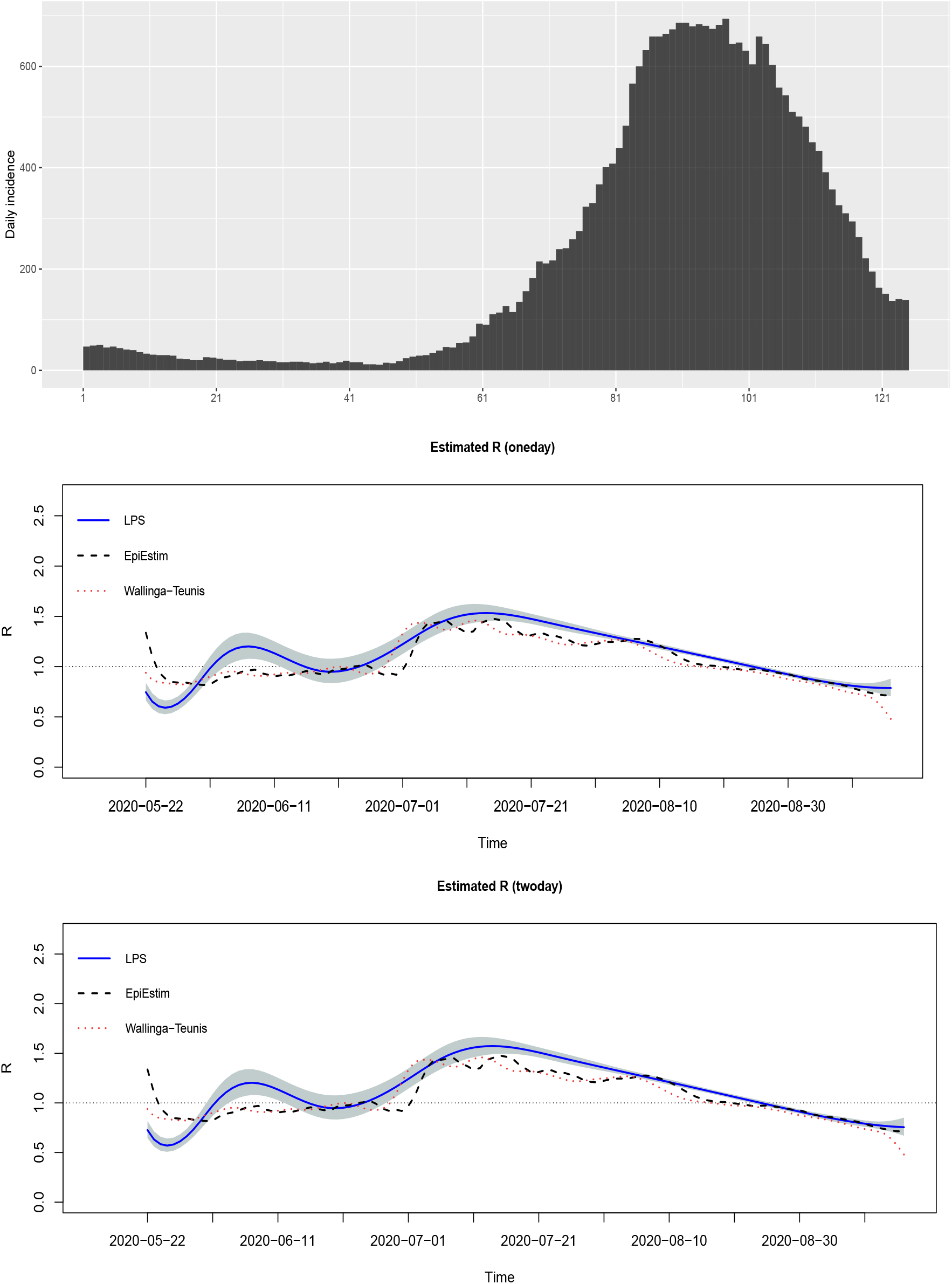
Daily incidence (top) and estimated *R*_*t*_ of Covid-19 for Australia between May and September 2020 with the **EpiEstim** (dashed), LPS (solid) and Wallinga-Teunis (dotted) method.

## 5 Discussion

The Laplacian-P-splines methodology presented in this paper combines Laplace approximation and Bayesian penalized B-splines for fast and flexible estimation of the time-varying reproduction number in an epidemic model with misreported data. The key benefit of our approach is its computational speed. While classic MCMC methods may take hours to deliver posterior estimates of key epidemiological parameters, estimation with LPS typically requires a couple of seconds.

This article shows that LPS performs at least as good as existing methods for real-time estimation of *R*_*t*_ such as **EpiEstim** or the Wallinga-Teunis approach. Moreover, it allows for different specifications of the delay pattern (one-day, two-day or weekend delays), covering practical scenarios arising in the real world during epidemic outbreaks. From here, several directions can be explored in the future to further improve the LPS methodology in the framework of epidemic modeling. First, it would be important to go beyond a naive multiplication factor approach to approximate the latent number of daily cases *M*_*t*_. This will probably improve the accuracy of posterior estimates for *R*_*t*_ and for the reporting and delay probabilities. Second, instead of using the maximum a posteriori (MAP) estimator for the hyperparameters, an alternative (and also more costly) strategy would be to use grid-based or MCMC approaches that would capture the uncertainty of posterior estimates more precisely and less locally than the MAP method considered here. Third, it would be interesting to refine the delay patterns and work for instance with ad hoc reporting structures that take into account public holidays. Finally, in the long term, we plan to make the routines of LPS available in a software package and hence create a simple and intuitive tool for fast approximate Bayesian inference in epidemic models with misreported data.

## Data Availability

Real data applications in Section 4 are publicly available. The links are provided below.

https://cran.r-project.org/package=COVID19

https://cran.r-project.org/package=EpiEstim

## Conflict of interest

The authors declare no conflicts of interest.

## Acknowledgments

This project is funded by the European Union’s Research and Innovation Action under the H2020 work programme, EpiPose (grant number 101003688).

## Appendix A

**Composition matrix for a one-day delay**

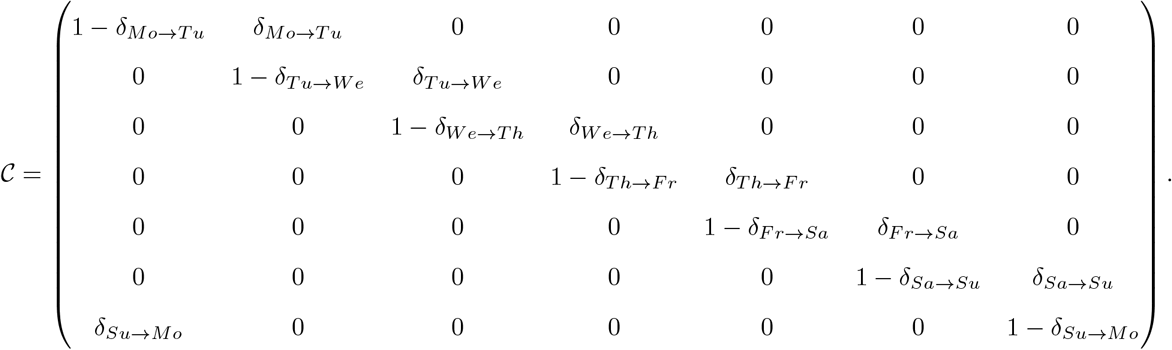

**Composition matrix for a two-day delay**

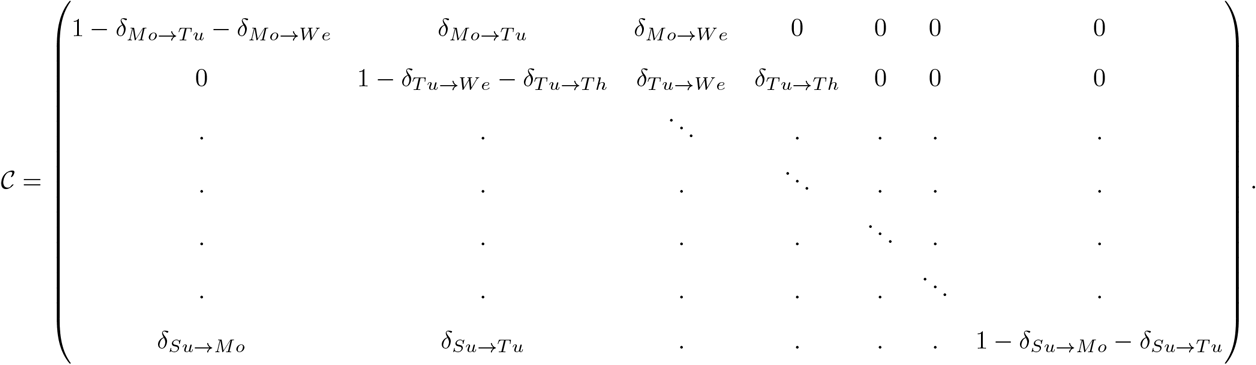

**Composition matrix for a weekend delay**

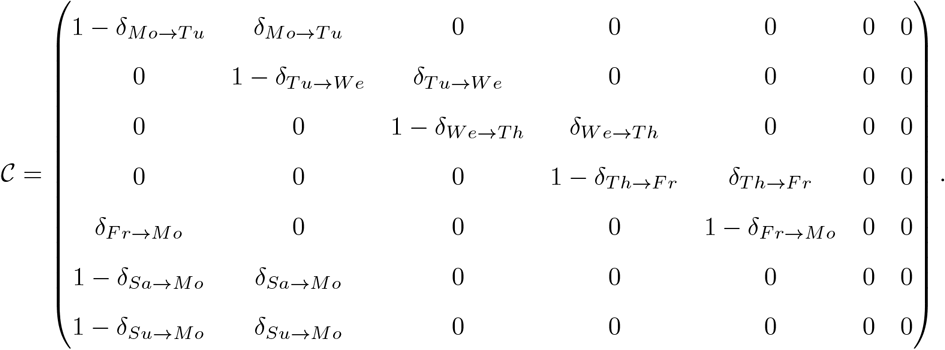

## Appendix B

The second-order Taylor expansion of *g*_*t*_(***θ, η***) around an initial vector ***θ***^(0)^ is written as:

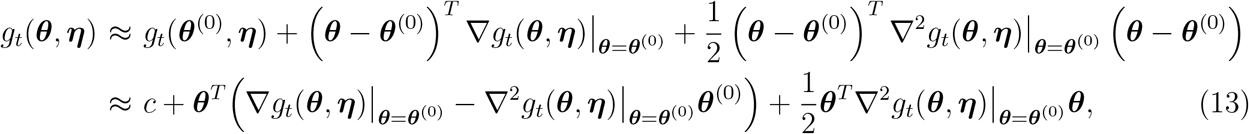

where *c* is a constant that does not depend on ***θ***. An analytical version of (13) is obtained by computing the following gradient and Hessian matrix:

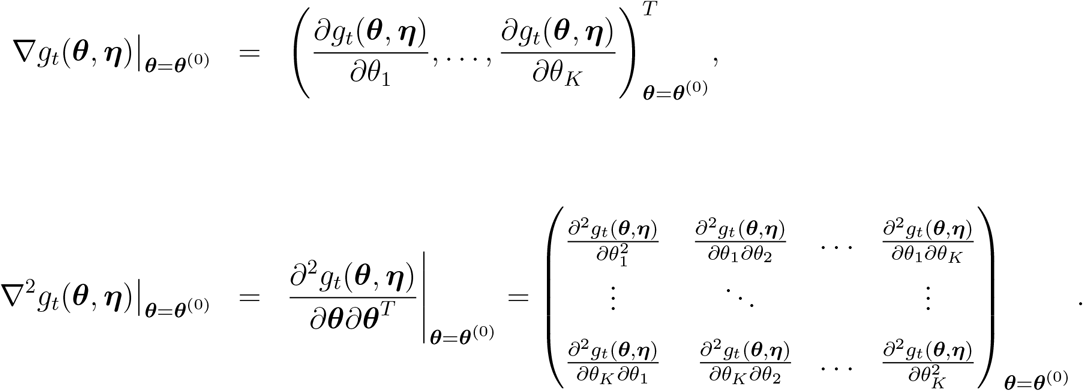

### Gradient

Recall that *g*_*t*_(***θ, η***) = *O*_*t*_ log(*s*_*t*_(***θ, η***)) − *s*_*t*_(***θ, η***), so the derivative with respect to the *k*th B-spline coefficient is:

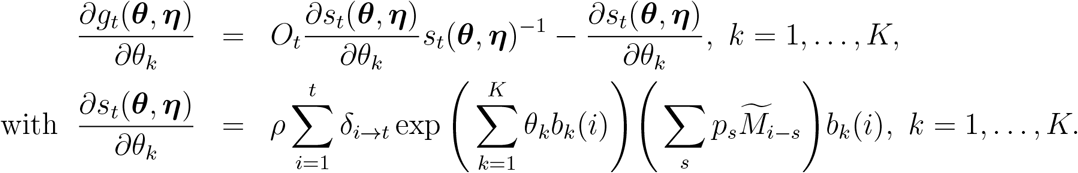

### Hessian

To obtain the *K* × *K* Hessian matrix, the following second-order partial derivatives for *k, l* = 1, …, *K* are computed:

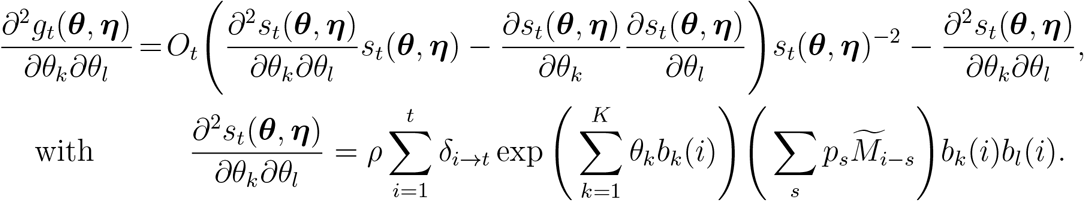

Having computed the gradient and Hessian for all day indexes of the epidemic *t* = 1, …, *T*, the results are summed up to compute the gradient and Hessian of the log-likelihood (across all observations), namely 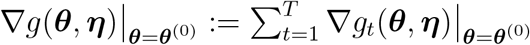 and 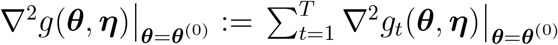. Using the second-order Taylor expansion in (13) (omitting the constant) and the log-likelihood function, we find:

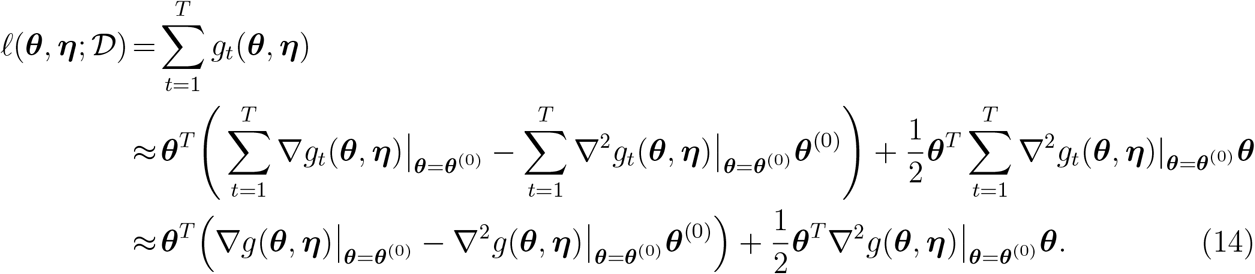

Plugging (14) in (6) and rearranging the terms, one gets the Laplace approximation to the conditional posterior of the vector of B-spline parameters:

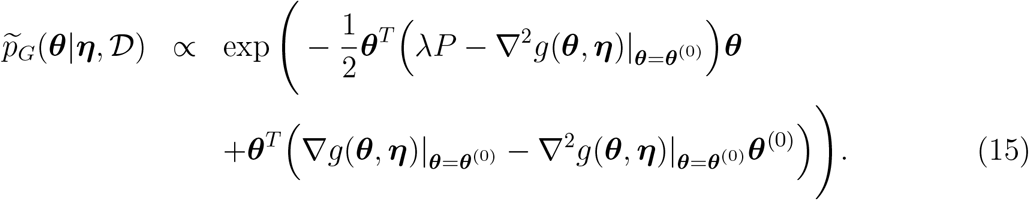

Note that (15) is (up to a multiplicative constant) a Gaussian density with mean (mode) and variance-covariance matrix equal to:

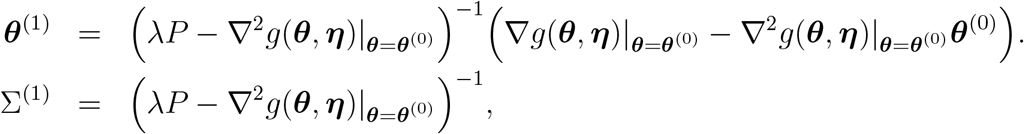

where the mean (mode) is obtained by solving the equation 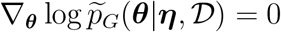 for ***θ*** and the variance-covariance matrix is 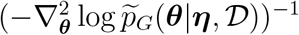. Let ***θ***\*(***η***) and Σ^*^(***η***) denote the mode and variance-covariance matrix towards which the iterative Laplace approximation scheme for *p*(***θ***|***η***, 𝒟) has converged for a given vector of hyperparameters ***η***. The final Laplace approximation is written (by abuse of notation) as:

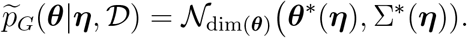

## Notes

### Competing Interest Statement

The authors have declared no competing interest.

## References

Arroyo-Marioli, F., Bullano, F., Kucinskas, S., and Rondón-Moreno, C. (2021). Tracking R of COVID-19: A new real-time estimation using the Kalman filter. PLOS One, 16(1):e0244474. https://doi.org/10.1371/journal.pone.0244474.

Azmon, A., Faes, C., and Hens, N. (2014). On the estimation of the reproduction number based on misreported epidemic data. Statistics in Medicine, 33(7):1176–1192. https://doi.org/10.1002/sim.6015.

Bettencourt, L. M. and Ribeiro, R. M. (2008). Real time Bayesian estimation of the epidemic potential of emerging infectious diseases. PLOS One, 3(5):e2185. https://doi.org/10.1371/journal.pone.0002185.

Bracher, J. and Held, L. (2020). A marginal moment matching approach for fitting endemic-epidemic models to underreported disease surveillance counts. Biometrics, pages 1–13. https://doi.org/10.1111/biom.13371.

Cauchemez, S., Boëlle, P.-Y., Thomas, G., and Valleron, A.-J. (2006). Estimating in real time the efficacy of measures to control emerging communicable diseases. American Journal of Epidemiology, 164(6):591–597. https://doi.org/10.1093/aje/kwj274.

Cori, A., Ferguson, N. M., Fraser, C., and Cauchemez, S. (2013). A new framework and software to estimate time-varying reproduction numbers during epidemics. American Journal of Epidemiology, 178(9):1505–1512. https://doi.org/10.1093/aje/kwt133.

Cui, J. and Kaldor, J. (1998). Changing pattern of delays in reporting AIDS diagnoses in Australia. Australian and New Zealand Journal of Public Health, 22(4):432–435. https://doi.org/10.1111/j.1467-842X.1998.tb01409.x.

Eilers, P. H. (2007). Ill-posed problems with counts, the composite link model and penalized likelihood. Statistical Modelling, 7(3):239–254. https://doi.org/10.1177%2F1471082X0700700302.

Eilers, P. H. C. and Marx, B. D. (1996). Flexible smoothing with B-splines and penalties. Statistical Science, 11(2):89–102. https://doi.org/10.1214/ss/1038425655.

Feller, W. (1941). On the integral equation of renewal theory. The Annals of Mathematical Statistics, 12(3):243–267. https://doi.org/10.1214/aoms/1177731708.

Fraser, C. (2007). Estimating individual and household reproduction numbers in an emerging epidemic. PLOS One, 2(8):e758. https://doi.org/10.1371/journal.pone. 0000758.

Fraser, C., Donnelly, C. A., Cauchemez, S., Hanage, W. P., Van Kerkhove, M. D., Hollingsworth, T. D., Griffin, J., Baggaley, R. F., Jenkins, H. E., Lyons, E. J., et al. (2009). Pandemic potential of a strain of influenza A (H1N1): Early findings. Science, 324(5934):1557–1561. https://doi.org/10.1126/science.1176062.

Goldfeld, S. M., Quandt, R. E., and Trotter, H. F. (1966). Maximization by quadratic hill-climbing. Econometrica: Journal of the Econometric Society, 34(3):541–551. https://doi.org/10.2307/1909768.

Gómez-Rubio, V. and Rue, H. (2018). Markov chain Monte Carlo with the Integrated Nested Laplace Approximation. Statistics and Computing, 28:1033–1051. https://doi.org/10.1007/s11222-017-9778-y.

Gostic, K. M., McGough, L., Baskerville, E. B., Abbott, S., Joshi, K., Tedijanto, C., Kahn, R., Niehus, R., Hay, J. A., De Salazar, P. M., et al. (2020). Practical considerations for measuring the effective reproductive number, Rt. PLOS Computational Biology, 16(12):e1008409. https://doi.org/10.1371/journal.pcbi.1008409.

Gressani, O. and Lambert, P. (2018). Fast Bayesian inference using Laplace approximations in a flexible promotion time cure model based on P-splines. Computational Statistics & Data Analysis, 124:151–167. https://doi.org/10.1016/j.csda.2018.02.007.

Gressani, O. and Lambert, P. (2021). Laplace approximations for fast Bayesian inference in generalized additive models based on P-splines. Computational Statistics & Data Analysis, 154:107088. https://doi.org/10.1016/j.csda.2020.107088.

Guidotti, E. and Ardia, D. (2020). Covid-19 Data Hub. Journal of Open Source Software, 5(51):2376. https://doi.org/10.21105/joss.02376.

Heesterbeek, H., Anderson, R. M., Andreasen, V., Bansal, S., De Angelis, D., Dye, C., Eames, K. T. D., Edmunds, W. J., Frost, S. D. W., Funk, S., Hollingsworth, T. D., House, T., Isham, V., Klepac, P., Lessler, J., Lloyd-Smith, J. O., Metcalf, C. J. E., Mollison, D., Pellis, L., Pulliam, J. R. C., Roberts, M. G., and Viboud, C. (2015). Modeling infectious disease dynamics in the complex landscape of global health. Science, 347(6227). https://doi.org/10.1126/science.aaa4339.

Hens, N., Van Ranst, M., Aerts, M., Robesyn, E., Van Damme, P., and Beutels, P. (2011). Estimating the effective reproduction number for pandemic influenza from notification data made publicly available in real time: a multi-country analysis for influenza A/H1N1v 2009. Vaccine, 29(5):896–904. https://doi.org/10.1016/j.vaccine.2010.05.010.

Hethcote, H. W. (2000). The mathematics of infectious diseases. SIAM Review, 42(4):599– 653. https://doi.org/10.1137/S0036144500371907.

Jandarov, R., Haran, M., Bjørnstad, O., and Grenfell, B. (2014). Emulating a gravity model to infer the spatiotemporal dynamics of an infectious disease. Journal of the Royal Statistical Society. Series C (Applied Statistics), 63(3):423–444. https://www.jstor.org/stable/24771855.

Lambert, P. and Eilers, P. H. (2005). Bayesian proportional hazards model with timevarying regression coefficients: a penalized Poisson regression approach. Statistics in Medicine, 24(24):3977–3989. https://doi.org/10.1002/sim.2396.

Lang, S. and Brezger, A. (2004). Bayesian P-splines. Journal of Computational and Graphical Statistics, 13(1):183–212. https://doi.org/10.1198/1061860043010.

Lawless, J. (1994). Adjustments for reporting delays and the prediction of occurred but not reported events. Canadian Journal of Statistics, 22(1):15–31. https://doi.org/10.2307/3315826.n1.

Levenberg, K. (1944). A method for the solution of certain non-linear problems in least squares. Quarterly of Applied Mathematics, 2(2):164–168. https://www.jstor.org/stable/43633451.

Lunn, D. J., Thomas, A., Best, N., and Spiegelhalter, D. (2000). WinBUGS - A Bayesian modelling framework: concepts, structure, and extensibility. Statistics and Computing, 10(4):325–337. https://doi.org/10.1023/A:1008929526011.

Marquardt, D. W. (1963). An algorithm for least-squares estimation of nonlinear parameters. Journal of the Society for Industrial and Applied Mathematics, 11(2):431–441. https://www.jstor.org/stable/2098941.

Martins, T. G. and Rue, H. (2014). Extending Integrated Nested Laplace approximation to a class of near-Gaussian latent models. Scandinavian Journal of Statistics, 41(4):893–912. https://doi.org/10.1111/sjos.12073.

Nishiura, H. and Chowell, G. (2009). The effective reproduction number as a prelude to statistical estimation of time-dependent epidemic trends. In Mathematical and Statistical estimation approaches in Epidemiology, pages 103–121. Springer. https://doi.org/10.1007/978-90-481-2313-1_5.

Nouvellet, P., Cori, A., Garske, T., Blake, I. M., Dorigatti, I., Hinsley, W., Jombart, T., Mills, H. L., Nedjati-Gilani, G., Van Kerkhove, M. D., Fraser, C., Donnelly, C. A., Ferguson, N. M., and Riley, S. (2018). A simple approach to measure transmissibility and forecast incidence. Epidemics, 22:29–35. https://doi.org/10.1016/j.epidem.2017.02.012.

Plummer, M. et al.. (2003). JAGS: A program for analysis of Bayesian graphical models using Gibbs sampling. In Proceedings of the 3rd international workshop on distributed Statistical Computing. Vienna, Austria. http://www.r-project.org/conferences/DSC-2003/.

Riou, J., Poletto, C., and Boëlle, P.-Y. (2018). Improving early epidemiological assessment of emerging aedes-transmitted epidemics using historical data. PLOS neglected tropical diseases, 12(6):e0006526. https://doi.org/10.1371/journal.pntd.0006526.

Rue, H., Martino, S., and Chopin, N. (2009). Approximate Bayesian inference for latent Gaussian models by using Integrated Nested Laplace approximations. Journal of the Royal Statistical Society: Series B (Statistical Methodology), 71(2):319–392. https://doi.org/10.1111/j.1467-9868.2008.00700.x.

Stocks, T., Britton, T., and Höhle, M. (2020). Model selection and parameter estimation for dynamic epidemic models via iterated filtering: application to rotavirus in Germany. Biostatistics, 21(3):400–416. https://doi.org/10.1093/biostatistics/kxy057.

Svensson, Å. (2007). A note on generation times in epidemic models. Mathematical Biosciences, 208(1):300–311. https://doi.org/10.1016/j.mbs.2006.10.010.

Thompson, R. and Baker, R. (1981). Composite link functions in generalized linear models. Journal of the Royal Statistical Society: Series C (Applied Statistics), 30(2):125–131. https://doi.org/10.2307/2346381.

Thompson, R., Stockwin, J., van Gaalen, R., Polonsky, J., Kamvar, Z., Demarsh, P., Dahlqwist, E., Li, S., Miguel, E., Jombart, T., Lessler, J., Cauchemez, S., and Cori, A. (2019). Improved inference of time-varying reproduction numbers during infectious disease outbreaks. Epidemics, 29:100356. https://doi.org/10.1016/j.epidem.2019.100356.

Tierney, L. and Kadane, J. B. (1986). Accurate approximations for posterior moments and marginal densities. Journal of the American Statistical Association, 81(393):82–86. https://www.tandfonline.com/doi/abs/10.1080/01621459.1986.10478240.

Vanhatalo, J., Riihimäki, J., Hartikainen, J., Jylänki, P., Tolvanen, V., and Vehtari, A. (2013). Gpstuff: Bayesian modeling with Gaussian processes. Journal of Machine Learning Research, 14:1175–1179. https://arxiv.org/abs/1206.5754.

Wallinga, J. and Lipsitch, M. (2007). How generation intervals shape the relationship between growth rates and reproductive numbers. Proceedings of the Royal Society B: Biological Sciences, 274(1609):599–604. https://doi.org/10.1098/rspb.2006.3754.

Wallinga, J. and Teunis, P. (2004). Different epidemic curves for severe acute respiratory syndrome reveal similar impacts of control measures. American Journal of Epidemiology, 160(6):509–516. https://doi.org/10.1093/aje/kwh255.

White, L. F. and Pagano, M. (2008). Transmissibility of the influenza virus in the 1918 pandemic. PLOS One, 3(1):e1498. https://doi.org/10.1371/journal.pone.0001498.

